# Radiological appearances of Anastomotic Leakage after Radical Gastrectomy

**DOI:** 10.1101/2020.04.25.20080093

**Authors:** Birendra Kumar Sah, Zhang Yang, Zhang Huan, Li Jian, Liu Wentao, Yan Chao, Li Chen, Yan Min, Zhu Zheng Gang

**Author notes:** Postal add: 197 Ruijin Er Road, Shanghai-200025, China, Contact no. (Co-first author).

## Abstract

**Background:** Anastomotic leakage is a critical postoperative complication after gastric cancer surgery. Previous studies have not specified radiological findings of anastomotic leakage. We investigated the potential burden caused by postoperative anastomotic leakage and explored the objective appearances of anastomotic leakage on computed tomography (CT) examination.

**Methods:** Gastric cancer patients who underwent curative gastrectomy and had a CT examination after surgery were included in this study. Propensity score (PS) matching generated 70 cases (35 cases of anastomotic leakage and 35 cases of no anastomotic leak) among 210 eligible cases. Univariate and multivariate analyses were used to identify the predictive variables of CT findings.

**Results:** More severe postoperative complications were observed in patients who had an anastomotic failure than those without anastomotic leakage(p<0.05). The median number of postoperative days (PODs) was 18 days for patients with no anastomotic leak, but the length of stay was almost three times longer (50 days) in patients with anastomotic leakage(p<0.05). In the univariate analysis, we observed a significant association between anastomotic leakage and five CT variables, including pneumoperitoneum, pneumoseroperitoneum (intra-abdominal accumulation of mixed gas and fluid), accumulation of extraluminal gas at the anastomosis site, seroperitoneum and extraluminal fluid collection at the anastomosis site (p<0.05). The multivariate analysis of the CT parameters revealed that the accumulation of extraluminal gas at the anastomosis site is the independent diagnostic parameters of a postoperative anastomotic leakage (p<0.05).

**Conclusions:** The occurrence of an anastomotic leakage significantly compromises the patients and increases the treatment burden. The CT variables of this study are beneficial to rule out anastomotic leakage after gastric cancer surgery. Extraluminal gas at the anastomosis site is highly suggestive of anastomotic leakage.

## Background

Postoperative complications were quite prevalent after gastric cancer surgery, and the complication rate was almost equal between open and laparoscopic surgery(1). However, severe complications occurred at a lower rate at high-volume centers than in low-volume centers (2). Generally, the rate of anastomotic leakage has been reported to be below five percent, or even below two percent in the experienced centers of many Asian countries (1, 3). Nevertheless, anastomotic leaks are still considered severe postoperative complications that aggravate the condition of compromised patients, and the mortality rate of patients with anastomotic leakages is significantly higher than that of patients without anastomotic leakages (4-6). Several scientific reports have explored predicting the postoperative complications of gastric cancer surgery; however, most of these reports were observational studies, and many risk factors were unavoidable in general practice (7-12).

Previous studies have not produced a better understanding of the anastomotic leakage after gastric cancer surgery. Computed tomography (CT) scans have been used to detect anastomotic leaks in patients after esophagectomy, but very few studies have reported on gastric cancer surgery, and the routine use of CT has been controversial (13, 14). No definitive suggestions exist on whether a postoperative CT or abdominal X-ray with an oral contrast agent should be routinely performed for the early detection of anastomotic leakage (15, 16). Furthermore, the interpretation of CT findings is highly subjective; for example, clinicians and radiologists are still unsure whether the presence of free gas in the abdominal cavity or the abdominal wall is normal after gastrectomy, whether this free gas is common after laparoscopic surgeries. Therefore, we conducted this study to explore the objective findings of the anastomotic leakage on computed tomography (CT).

## Methods

This is a retrospective study, and the primary inclusion criterion was gastric cancer patients who underwent curative gastrectomy and a CT examination after surgery. The study endpoint was the presence of postoperative complications and any complications within one month of discharge from the hospital. Altogether, 221 patients were identified, and 11 patients with benign diseases were excluded. Finally, we a total of 210 patients with gastric cancer diseases were included. All patients underwent curative gastrectomies with appropriate lymph node dissections between November 2015 and August 2018 and received a postoperative CT scan. Propensity score (PS) matching generated 70 eligible cases (35 cases of anastomotic leakage and 35 cases of no anastomotic leak) with five covariates, i.e., age, body mass index (BMI), mode of surgery (open or laparoscopic), extent of resection (subtotal or total), and combined resection of adjacent organs (Table 1).

**Table 1.** Demographic data of the PS-matched patients

| Parameter |  | Without leakage | Leakage |
| --- | --- | --- | --- |
| Sex | Male | 27 | 25 |
|  | Female | 8 | 10 |
| Age group (years) | 41–50 | 2 | 2 |
|  | 51–60 | 7 | 8 |
|  | 61–70 | 11 | 9 |
|  | 71–80 | 10 | 11 |
|  | 81–90 | 5 | 5 |
| BMI | <25 | 24 | 27 |
|  | 25–<30 | 11 | 7 |
|  | ≥30 | 0 | 1 |
| Mode of surgery | Open | 33 | 33 |
|  | Laparoscopic | 2 | 2 |
| Type of resection | Subtotal | 19 | 19 |
|  | Total | 16 | 16 |
| Combined resection | No | 28 | 29 |
|  | Yes | 7 | 6 |
| TNM Stage | I | 13 | 8 |
|  | II | 4 | 3 |
|  | III | 15 | 21 |

Two experienced radiologists with gastrointestinal expertise carefully reviewed all CT scans. Both radiologists were provided with clinical information, including the surgery mode and anastomosis type, but were blinded to the clinical results regarding anastomotic leakage. Any disagreements about the CT findings were settled by discussion between the radiologists for and a final consensus. We collected detailed clinical parameters, including vital signs, blood test results, surgery type, and the TNM classification of the tumour. The treating doctors recorded thirty-five cases of anastomotic leakage.

One of the following observations was required for the diagnosis of an anastomotic leakage: 1. confirmation by reoperation; 2. presence of digestive content, food debris or methylene blue in the abdominal drainage tube; and 3. clear images of extraluminal contrast leak on the CT scan.

### Statistical analysis

The Statistical Package for Social Science (SPSS) version 22.0 for Windows (SPSS, Inc., Chicago, Illinois) was applied for the statistical analysis. Nonparametric methods were used to test data with an abnormal distribution. A chi-square test or Fisher’s exact test was used to compare the differences between the two groups. Logistic regression was applied to identify the independent predictive factors for anastomotic leakage. A p-value of less than 0.05 was considered statistically significant.

## Results

There was no difference in the basic clinical parameters between the two groups (p<0.05, Table 1). The reoperation rate was significantly higher in patients with anastomotic leakage than in those without an anastomotic leakage. More hemorrhagic, infectious complications, and impaired vital organ function were observed in patients who had an anastomotic failure than those without anastomotic leakage (p<0.05, Table 2). Four patients died of postoperative complications after developing an anastomotic leakage. The leading cause of death was severe abdominal infection followed by shock and cardiac and respiratory failure. There were no deaths in the group without anastomotic leakages.

**Table 2.** Postoperative complications between the two groups

| Complication |  | Number of patients |  | p-value |
| --- | --- | --- | --- | --- |
|  |  | Without leakage | With leakage |  |
| Hemorrhage | Intra-abdominal | 0 | 6 | 0.025 |
|  | Anastomosis site | 0 | 2 | NS |
| Wound dehiscence |  | 2 | 6 | NS |
| Multiple infections |  | 2 | 13 | 0.003 |
| Overall infection |  | 17 | 25 | 0.051 |
| Impaired renal function |  | 4 | 14 | 0.006 |
| Cardiac/respiratory failure |  | 2 | 10 | 0.023 |
| Overall gastrointestinal obstruction |  | 8 | 13 | NS |
| Pancreatic fistula |  | 1 | 0 | NS |
| Readmission |  | 1 | 3 | NS |
| Reoperation |  | 2 | 10 | 0.023 |
| Death |  | 0 | 4 | NS |

The postoperative length of hospital stay was significantly longer for patients with an anastomotic leakage than for those without anastomotic leakage (p<0.05); the median number of postoperative days (PODs) was 18 days for patients with no anastomotic leak, but the length of stay was almost three times longer (50 days) in patients with anastomotic leakage. The overall expenditure was significantly different between the two groups (p<0.05). The median total expenditure for patients with no leakage was only 64193.46 RMB (Chinese currency), but the expenditure was almost double (121167.12 RMB) for patients who had an anastomotic leakage.

In the univariate analysis, we observed a significant association between anastomotic leakage and five CT variables, including pneumoperitoneum, pneumoseroperitoneum (intra-abdominal accumulation of mixed gas and fluid), accumulation of extraluminal gas at the anastomosis site, seroperitoneum and extraluminal fluid collection at the anastomosis site (Table 3, p<0.05). The multivariate analysis of the CT parameters revealed that the accumulation of extraluminal gas at the anastomosis site is the independent diagnostic parameters of a postoperative anastomotic leakage (p<0.05, Odds ratio 5.88, 95% CI 1.84–18.83). About 78.6 percent of patients with extraluminal gas at the anastomosis site were diagnosed of having anastomotic leakage. Typical images of accumulation of extraluminal gas at different types of anastomosis sites were visible in CT scan (Fig.1, 2, 3).

**Fig. 1.**
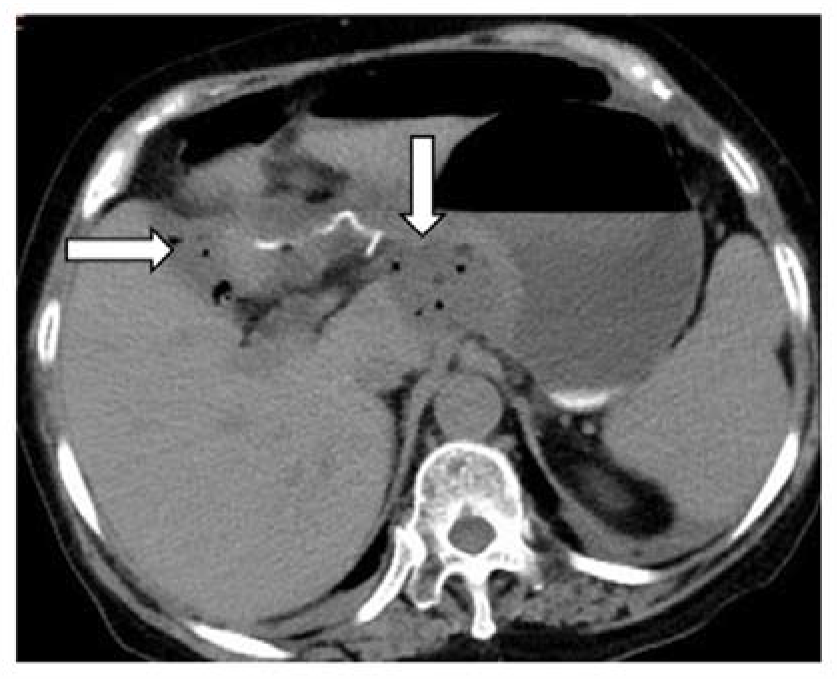
Extraluminal gas at gastroduodenal anastomosis

**Fig. 2.**
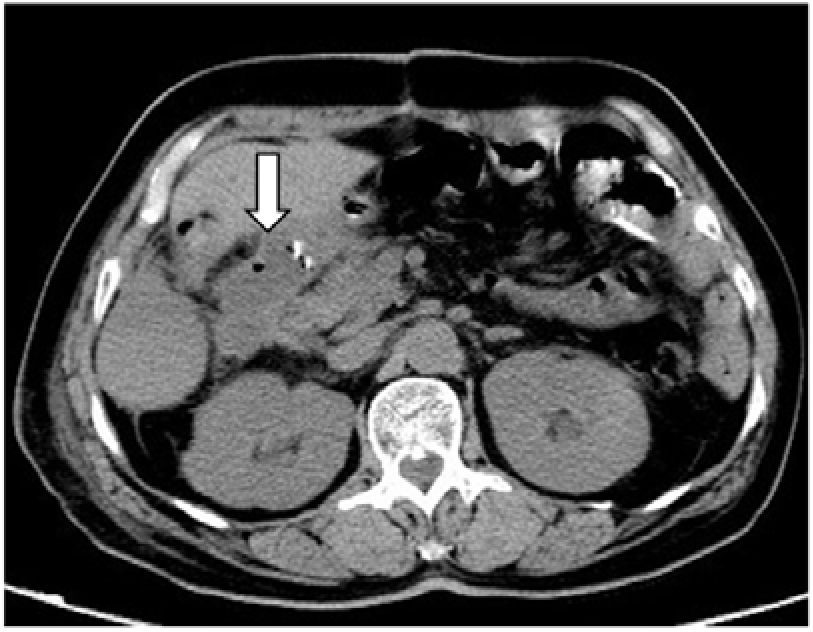
Extraluminal gas at duodenal stump

**Fig. 3.**
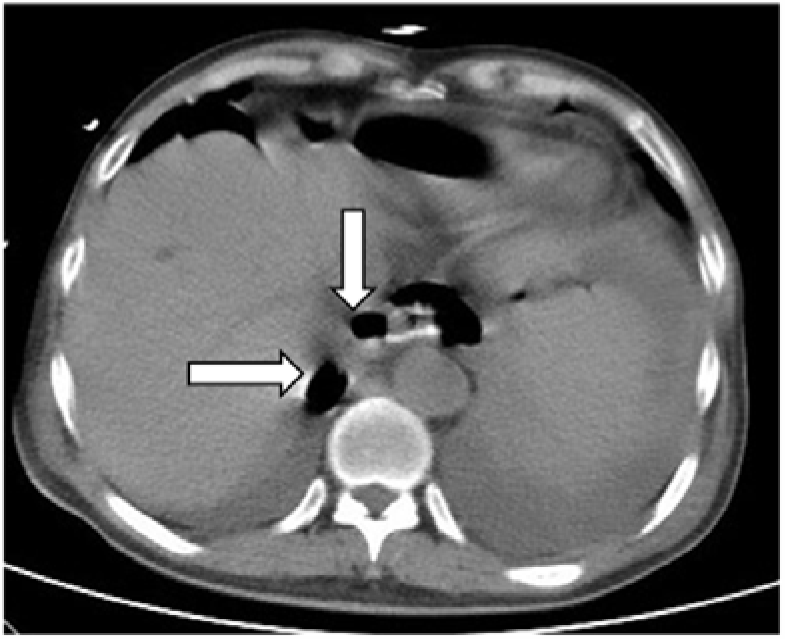
Extraluminal gas at esophagojejunal anastomosis

**Table 3.** Univariate analysis of the postoperative CT findings

| Factor |  | Without leakage | With leakage | p value |
| --- | --- | --- | --- | --- |
| Pneumoperitoneum | Absent | 17 (70.8) | 7 (29.2) | 0.012 |
|  | Present | 18 (39.1) | 28 (60.9) |  |
| Anastomosis site gas | Absent | 29 (69.0) | 13 (31.0) | 0.000 |
|  | Present | 6 (21.4) | 22 (78.6) |  |
| Seroperitoneum | Absent | 15 (83.3) | 3 (16.7) | 0.002 |
|  | Present | 20 (38.5) | 32 (61.5) |  |
| Anastomosis site fluid | Absent | 26 (59.1) | 18 (40.9) | 0.048 |
|  | Present | 9 (34.6) | 17 (65.4) |  |
| Abdominal gas and fluid | Absent | 29 (65.9) | 15 (34.1) | 0.001 |
|  | Present | 6 (23.1) | 20 (76.9) |  |
| Abdominal wall gas | Absent | 29 (51.8) | 27 (48.2) | NS |
|  | Present | 6 (42.9) | 8 (57.1) |  |

## Discussion

For the analysis of any clinical condition with a low prevalence rate, a major hurdle is the statistical calculation. The analysis needs a substantial cohort to obtain statistically significant results. An anastomotic leakage rate of two percent corresponds to 98 out of 100 patients having a satisfactory recovery, regardless of the scenario. Therefore, we applied PS matching to standardize the data and facilitate a better comparison of clinical conditions between two groups. Many previous conventional studies have suggested that postoperative complications might be related to age, obesity, mode of surgery, and the extent of resection (7, 9, 10). Therefore, we incorporated all these factors as covariates for PS matching.

Many authors have advocated for barium swallow tests to diagnose suspicious cases of anastomotic leakages after gastrointestinal (GI) surgeries. Nevertheless, the generalized use of this examination is debatable (17-19). Few studies have suggested postoperative CT after gastric cancer surgery, and there are different opinions on the use of oral contrast agents (16, 20). We found there was no apparent benefit of using oral contrast agents to detect anastomotic leakages, as none of the 27 patients who ingested an oral contrast agent 6 hours before the CT scan had an extraluminal contrast leak. Besides,there are still many unanswered questions, for instance, how much oral contrast agent is needed, what is the optimal concentration, what is the optimal timing to orally ingest the contrast agent? These questions warrant a well-controlled future study regarding whether oral contrast agents are beneficial for diagnosing anastomotic leakages.

In this study, the univariate analysis found that five CT variables were significantly correlated with an anastomotic leak. These variables were created to minimize the subjective nature of having radiologists and surgeons judge the scans so that objective analyses can be applied in future clinical work. However, we still can not fully depend on CT examinations, as we did not find any CT sign with a specificity of 100 percent. Approximately 39 percent of patients with intra-abdominal free gas had no anastomotic leakage. Even the accumulation of gas at the anastomotic site led to an approximately 21 percent false-positive rate. Nevertheless, the results of this study identified some significant findings from the postoperative CT examinations, which were either unknown or not well-described in previous publications.

## Conclusion

The occurrence of an anastomotic leakage significantly compromises the patients and increases the treatment burden. The suggested CT findings from this study are beneficial to rule out anastomotic leakage after gastric cancer surgery. Extraluminal gas at the anastomosis site is highly suggestive of anastomotic leakage.

## Data Availability

The datasets used and/or analyzed during the current study are available from the corresponding author on reasonable request.

